# THE EFFECTIVENESS OF ACB-IP 1.0 UNIVERSAL PATHOGEN FREE CONCENTRATED COCKTAIL CONVALESCENT PLASMA IN COVID-19 INFECTION

**DOI:** 10.1101/2021.03.05.21251413

**Authors:** Cansu Hemsinlioglu, Nil Banu Pelit, Koray Yalcin, Omur Selin Gunaydın, Nihal Ozturk Sahin, Esra Savas Karagacli, Omer Elibol, Sefa Onur Demir, Evren Safak, Raife Dilek Turan, Goncagul Celebi, Miyase Ezgi Kocaoglu, Gozde Sir Karakus, Bulut Yurtsever, Cihan Tastan, Selen Abanuz, Didem Cakirsoy, Derya Dilek Kancagi, Zeynep Torun, Utku Seyis, Muhammer Elek, Rehile Zengin, Ayse Sesin Kocagoz, Caglar Cuhadaroglu, Nur Birgen, Siret Ratip, Ercument Ovali

**Affiliations:** Acıbadem Labcell Cellular Therapy Center, Istanbul; Acibadem Labmed Blood Banks, Istanbul; Acibadem Altunizade Hospital, Apheresis Center, Istanbul; Acibadem Altunizade Hospital Infectious Disease Unit, Istanbul; Acibadem Altunizade Hospital Intensive Care Unit, Istanbul; Acıbadem Altunizade Hospital, Istanbul; Acibadem Altunizade Hospital Bone Marrow Transplantation Unit, Istanbul

**Author notes:** **Correspondence:** Cansu HEMSİNLİOGLU, Icerenkoy, Kerem Aydınlar Kampusu, Acibadem Labcell, Kayısdagı Cd. No:32, 34752 Atasehir/Istanbul, Turkey.

**Keywords:** convalescent plasma, SARS-CoV2, COVID-19, neutralizing antibody

## Abstract

**Introduction:** The efficacy of SARS-CoV2 standard single donor convalescent plasma varied according to the application time and most importantly the amount of antibody that is administered. Single donor plasma has some drawbacks; such as the insufficient levels of neutralizing antibody activities, the requirements of blood group compatibility, and the risk of infection transmission. In this study, the efficacy and safety of pathogen inactivated, isohemagglutinin-depleted (concentrated) and pooled convalescent plasma was investigated.

**Methods:** In this study, ACB-IP 1.0 convalescent plasma product was prepared as follows; first, convalescent plasma was collected from different donors, then pathogen-inactivation was carried-out, and isohemagglutinins were cryodepleted, respectively. Finally, concentrated convalescent plasma product was pooled and stored until use.

A total of sixteen patients were treated with two different convalescent plasma products. Nine patients were treated with standard single donor convalescent plasma and seven were treated with pathogen-free, concentrated, pooled convalescent plasma (ACB-IP 1.0) between 01 April 2020 and 31 December 2020.

The outcomes of these two plasma products were compared regarding SARS-CoV2 antibody titers, neutralizing antibody activities, length of hospitalization and mortality rates.

**Results:** Five out of six single donor plasma SARS-CoV2 antibody titers remained below 12 s/co, but the antibody titers of all ACB-IP 1.0 plasma were above 12 s/co. SARS-CoV2 total antibody titers of ACB-IP 1.0 plasma were statistically higher than the antibody titers of single donor plasma. Mean total plasma neutralizing antibody activity of ACB-IP 1.0 plasma (1.5421) was found statistically higher than single donor plasma (0.9642) in 1:256 dilution (ρ=0.0087)

The mortality rate of the patients treated with ACB-IP 1.0 plasma showed statistically lower (p: 0,033) than the patients treated with single donor plasma. The administration of either single donor plasma or ACB-IP 1.0 plasma to the patients within eight days significantly shortened the length of hospitalization compared to administration of either plasma to the patients later than eight days (ρ= 0,0021)

**Discussion:** Pathogen-free, concentrated, pooled convalescent plasma may resolve the bias in SARS-CoV2 antibody titers and neutralizing antibody activities, without requiring blood group compatibility that allows patient accessibility in a shorter time and has safe plasma characteristic. This study indicates that ACB-IP 1.0 may be a superior product compared to standard single donor plasma.

(Patent Application No: PY2020-00232)

## INTRODUCTION

On 31 December 2019, a new case of pneumonia of anonymous etiology emerged in Wuhan City, Hubei Province China and humanity confronted a new pandemia. The World Health Organization (WHO) officially announced the causative organism as 2019-nCoV/SARS-CoV2.^1^ Viral genome sequence of this new human pathogen was released and found to be closely related to viral species called Severe Acute Respiratory Syndrome Coronavirus (SARS-CoV) which caused outbreaks in 2002 and 2003 in China.^2,3^

The SARS-CoV2 virus has spread from Wuhan to the whole of China and 223 countries worldwide, affected more than 101,561,219 individuals, resulted in over 2,196,944 deaths. Even though there are some vaccines available, there are still no monoclonal antibodies (mAbs) or drugs available for SARS-CoV2 infection^4^ due to numerous uncertainties about SARS-CoV2 infection such as viral replication kinetics, host interaction, immunomodulatory capacity, and its long term effects on individuals.^5^

Although many therapies are in a development phase, the safest alternative in clinical practice for immediate use appears to be human convalescent plasma. The fundamental mechanism behind convalescent plasma is passive antibody therapy for viral neutralization.^5^ Convalescent plasma containing antibody from SARS-CoV2 individuals who have recovered from the disease, has been suggested as an investigational treatment option by FDA.^6^

Our knowledge regarding convalescent human plasma comes from the treatment of viral infections, such as SARS-CoV, avian influenza A (H5N1) virus, influenza A (H1N1), MERS, and Ebola virus.^7-12^ Even though there is still no consensus about its effectiveness^5^, the efficacy of convalescent plasma varied according to the virus type, the application time, and most importantly the amount of antibody that is administered. There are similar concerns regarding the studies conducted in SARS-CoV2.^13, 14^

Single donor plasma has some drawbacks; such as the insufficient levels of neutralizing antibody titers^15-17^ the requirements of blood group compatibility^13, 16-18^, and the risk of infection (HIV, HBV, HCV… etc.) transmission by the donors who do not meet the standard donor criteria. In addition, excess of procoagulant factors in standard donor plasma also increases the risk of thrombosis. Therefore, a necessity for a new convalescent plasma product arose.

For this purpose, ACB-IP 1.0 cocktail plasma, which SARS-CoV2 antibody titers and neutralizing antibody activity tended to be standardized by pooling, is designed. Furthermore, Immunoglobin M (IgM) which is responsible for most of the isohemagglutinins^19-21^ are depleted by subjecting to cryodepletion process and, the plasma is pooled so that Anti-A and Anti-B isohemagglutinin titers were below 1/8. With the help of the cryodepletion process, the procoagulant factors are reduced so that risk of thrombotic events are tried to be eliminated. On the other hand, it is concentrated, and by subjecting it to pathogen inactivation, the safety of the plasma is improved.

Hence, the current study was carried out in 4 participating hospitals with 16 patients to compare both product quality and clinical effectiveness of ACB IP 1.0 and standard convalescent plasma.

## MATERIALS AND METHODS

According to the criteria specified in the COVID-19 Immune Plasma Procurement and Clinical Use Guidelines of the Ministry of Health in Turkey and FDA^6^, donor candidates who were eligible according to apheresis donor criteria were invited to the Acibadem Altunizade Hospital;Therapeutic Apheresis Center. Blood products were taken from the donor plasma candidates. (Supplementary-1)

Convalescent plasma samples, which were obtained from 9 Turkish Red Crescent donors and 7 ACB-IP 1.0 plasma prepared from mostly 8 (only 1 of the ACB-IP 1.0 plasma was prepared from 4 different donors) different donors were compared according to patients’ outcomes.

### Plasma Collection

ACB IP 1.0 obtained from the donor plasma using the TERUMO BCT Trima Accel device. 400 ml - 600 ml plasma was collected according to the patient’s height, weight, and hemogram results. During this process, an ISBT code was obtained from the Turkish Red Crescent.

### Pathogen Inactivation

Plasma collected by plasmapheresis were connected to Cerus Intercept Blood System INT 31 plasma treatment bags (Intercept Blood System INT 31 plasma treatment bags Lot No: CE19G18L71) using the bag joining device (Terumo TSCD-II TSCD Wafers Code No: SC*W017). Before the pathogen inactivation process, 2 ml of 2 tubes witness samples were taken from the collected plasma. Witness samples are stored at −40 °C. According to the manufacturer instructor, at first, plasma is treated with Amotosalen then photochemically irradiated with UVA at 320-400 nm wavelength in Intercept INT100 illuminator. After inactivation, the plasma is passed through the adsorption filter to remove unreactive amotosalen and free photoproducts, and then it is divided into 2 or 3 equal volumes, depending on the volume of the collected plasma. After this process, 2 ml of 2 tubes witness samples are taken and stored at −40 °C. The pathogen inactivated plasma is stored at −40 °C for labeling until the pooling process for clinical use.

### Isohemagglutinin Assay

Ready to use (commercial) A and B kits are provided by the Biorad and the gel centrifugation method was performed according to the manufacturer’s protocol^22^.

### Mini-pool Isohemagglutinin Depletion and Concentration of Plasma

After the plasma was obtained from the donor, Anti-A and Anti-B isohemagglutinin titers were determined. Management of Isohemagglutinin titer was carried out in two separate steps. In the first step, isohemagglutinins, most of which are of IgM nature, were reduced by cryodepletion, while simultaneously concentrating the product. In the second step, isohemagglutinin titer was tried to be reduced by pooling of Anti-A and Anti-B free plasma and the plasma containing them.

#### Cryodepletion Method

Plasma samples from the apheresis product of 200 ml were transferred to plasma storage bags (Terumo Flex transfer bag Lot No: 190912F2) frozen in a deep freezer at −40 °C. One bag consists of eight donors apheresis plasma and the volume is approximately 1600 ml. Frozen samples are kept at + 4 °C for defrosting for approximately 8-12 hours. Liquid plasma is separated from the cryoprecipitate by centrifugation. Witness samples are taken from the liquid plasma before separation and stored at −40 °C. The bag where the cryodepleted pool will be produced was connected to the plasma bag (Trima Accel Lot No: 1901175251) using the bag joining device (Terumo TSCD-II TSCD Wafers Code No: SC*W017).

#### The Pooling of Plasma

The plasma bag was placed in the extractor and the cryopoor plasma was transferred to the pooling bag. Finally, pooling was achieved, by mixing the low SARS-CoV2 antibody titer with high antibody titer plasma before cryodepletion and the total product is packaged in 200 ml bags and stored at −40 °C until use.

### SARS-CoV2 Specific Immunoglobulin Analysis

SARS-CoV2 specific immunoglobulin analyzes were performed using the CLIA method (Centaure XP Lot No: 005).

### Neutralizing Antibody Assay^**23**^

100 TCID50 / 50 microliter SARS-CoV2 virus is placed in 96 Well U Bottom plate and 50 µl diluted human serums (1:64, 1:128, 1:256 serum concentration) were added.

After one hour of incubation at room temperature, 10000 Vero cell/well is placed in a 96 Well Flat Bottom plate with 100 µl of complete DMEM (4% FBS + 1% PSA). Supernatants are removed after 72 hours of incubation, 50 μl of MTT solution and 50 μl serum-free media was added to the remaining cells. After incubating at 37 ° C for 4 hours, 100 μl Isopropanol dispersion is added to each well and placed on a shaker for 10 min. The results are obtained by ELISA Reader at an absorbent value of 570 nm. The neutralizing antibody activity was studied triplicated in 1:64, 1:128 and 1:256 dilution based by cell viability index.

### Endotoxin Analysis

The gel-clot technique is used for detecting or quantifying endotoxins (Division of Charles River Laboratories, Inc Lot: L4451L).

### Microbiological Quality Control

The pooled convalescent plasma is placed on to the Bactec Fx device for microbiological quality control analysis (Becton Dickinson).

### Sars-CoV2 Quantitative Real-Time Polymerase Chain Reaction (PCR) Test

After experienced healthcare provider had taken nasopharyngeal swap sample from COVID-19 positive for the qualitative detection of nucleic acid from SARS-CoV2 in upper respiratory specimens, the analysis was performed by using Quantitative Real-Time PCR Coronavirus Detection test kit according to the manufacturer’s instructions (Diacarta Lot No:2008711)

### Trial Design (NCT04769245)

A total of 16 Hospitalized adults were screened for enrollment and if they had positive reverse-transcriptase–polymerase-chain-reaction (RT-PCR) for SARS-CoV2 and radiologically confirmed pneumonia, were included in the study.

A total of sixteen patients were treated with two different convalescent plasma products. Nine patients were treated with single donor convalescent plasma and seven were treated with pathogen-free, concentrated, pooled convalescent plasma between 01 April 2020 and 31 December 2020.

Written informed consent was obtained from all patients or their first degree relatives, and the trial was conducted under the principles stated in the Declaration of Helsinki and Good Clinical Practice guidelines and approval of Acibadem University ethics committee (Approval No: 2020-06/02)

Clinical information of the groups was obtained from the hospital’s electronic medical records. Demographic data, presenting symptoms as well as a radiological presentation on the onset of disease including fever, cough, fatigue, dyspnea, diarrhea, oxygen requirement, treatments received (Ministry of Health of Turkey Covid-19 treatment algorithms), duration of hospitalization stay, duration of Intensive care unit (ICU) stay, cycles and volume of convalescent plasma received, symptom and radiological improvements, the current status of the patient were collected. These two groups were compared according to safety and efficacy. Patients were followed up for transfusion-related reactions and the findings were recorded.

### Statistical Analysis

SARS-CoV2 Antibody titers, neutralizing antibody activities, and duration of hospitalization were analyzed with Mann Whitney test. The mean age of the groups was compared by the Kolmogorov-Smirnov test. Survival differences between the two plasma groups were analyzed with the Chi-Square test. Moreover, Fisher exact test was used to compare the categorical variables such as radiological presentation, co-existing disease, and oxygen supplement requirement among the groups. Results were at a %95 confidence interval and a significant level of p=0.05 was used for all statistical analysis.

## RESULTS

### Preclinical Results

Analysis of the Isohemagglutinin Titers:

The analysis of the isohemagglutinin titers revealed that maximum Anti-A titers were; 1:64, Anti-B titers were 1:128 for single donor plasma and maximum Anti-A titers were; 1:4, Anti-B titers were 1:8 for ACB-IP 1.0.

#### Analysis of the SARS-CoV2 Antibody Titers in Plasma

Five out of six single donor plasma SARS-CoV2 antibody titers remained below 12 s/co, but the antibody titers of all the ACB-IP 1.0 plasma were above 12 s/co. The mean antibody titer of single donor plasma was measured as 9.2083, mean antibody titer of ACB-IP 1.0 plasma was measured as 32.700. SARS-CoV2 total antibody titers of ACB-IP 1.0 plasma were statistically higher than the antibody titers of single donor plasma (Figure 1).

**Figure 1:**
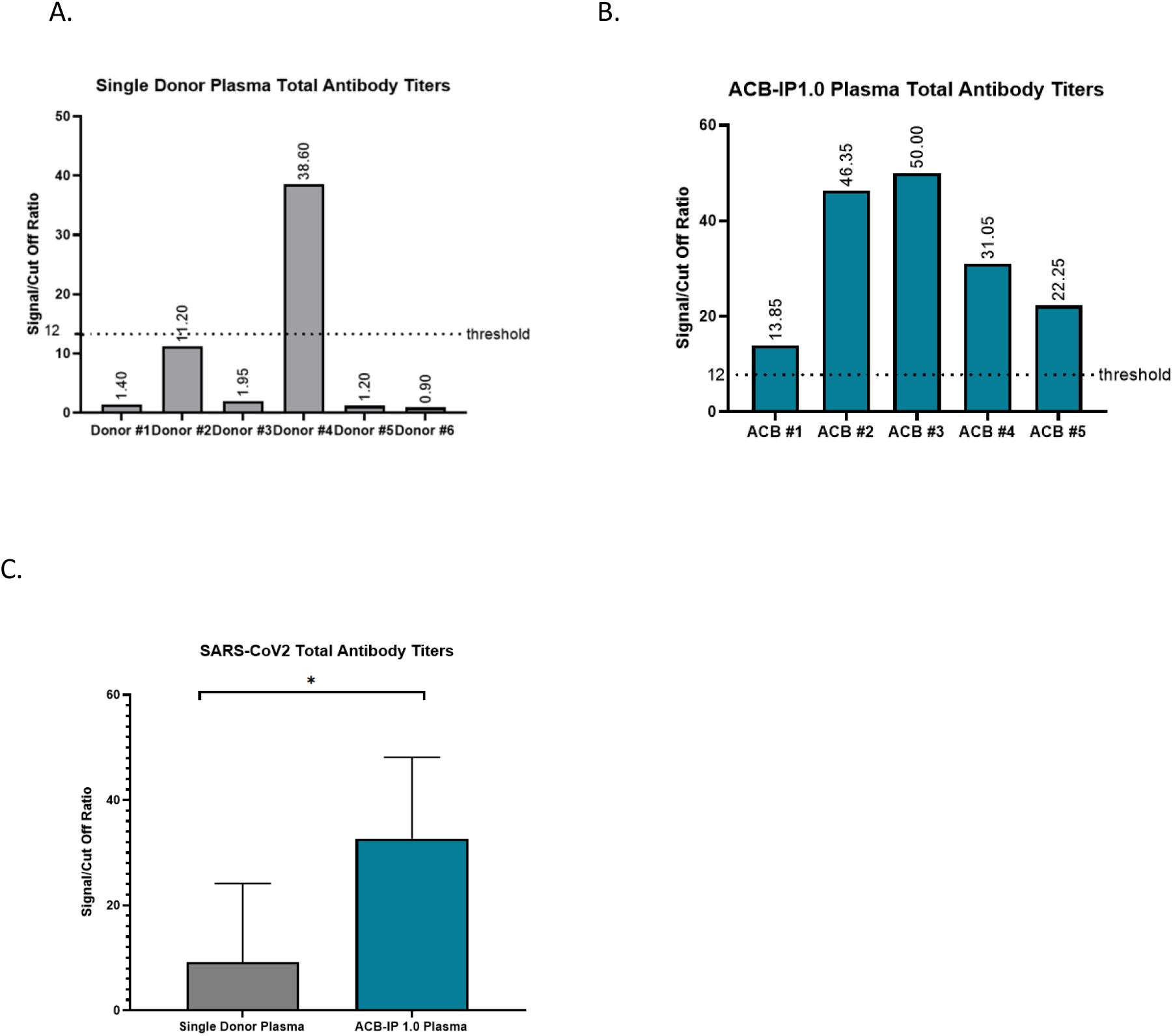
Single Donor Plasma and ACB-IP 1.0 Plasma SARS-CoV2 Antibody Titers A. Single donor plasma total antibody titers B.ACB-IP 1.0 plasma total antibody titers C. Comparison of mean antibody titers of single donor plasma and ACB-IP 1.0 plasma (* ρ=0.03)

#### Analysis of SARS-CoV2 Neutralizing Antibody Activity

Unlike expected, the SARS-CoV2 neutralizing capacity of the single donor plasma antibodies was higher than SARS-CoV2 antibody titers. Only 50% single donor plasma neutralizing antibody activity was below %100 cell viability threshold in 1:256 dilution (Figure 2). Whereas neutralizing activity of all ACB-IP1.0 plasma was above %100 cell viability threshold in 1:256 dilution (Figure 2).

**Figure 2:**
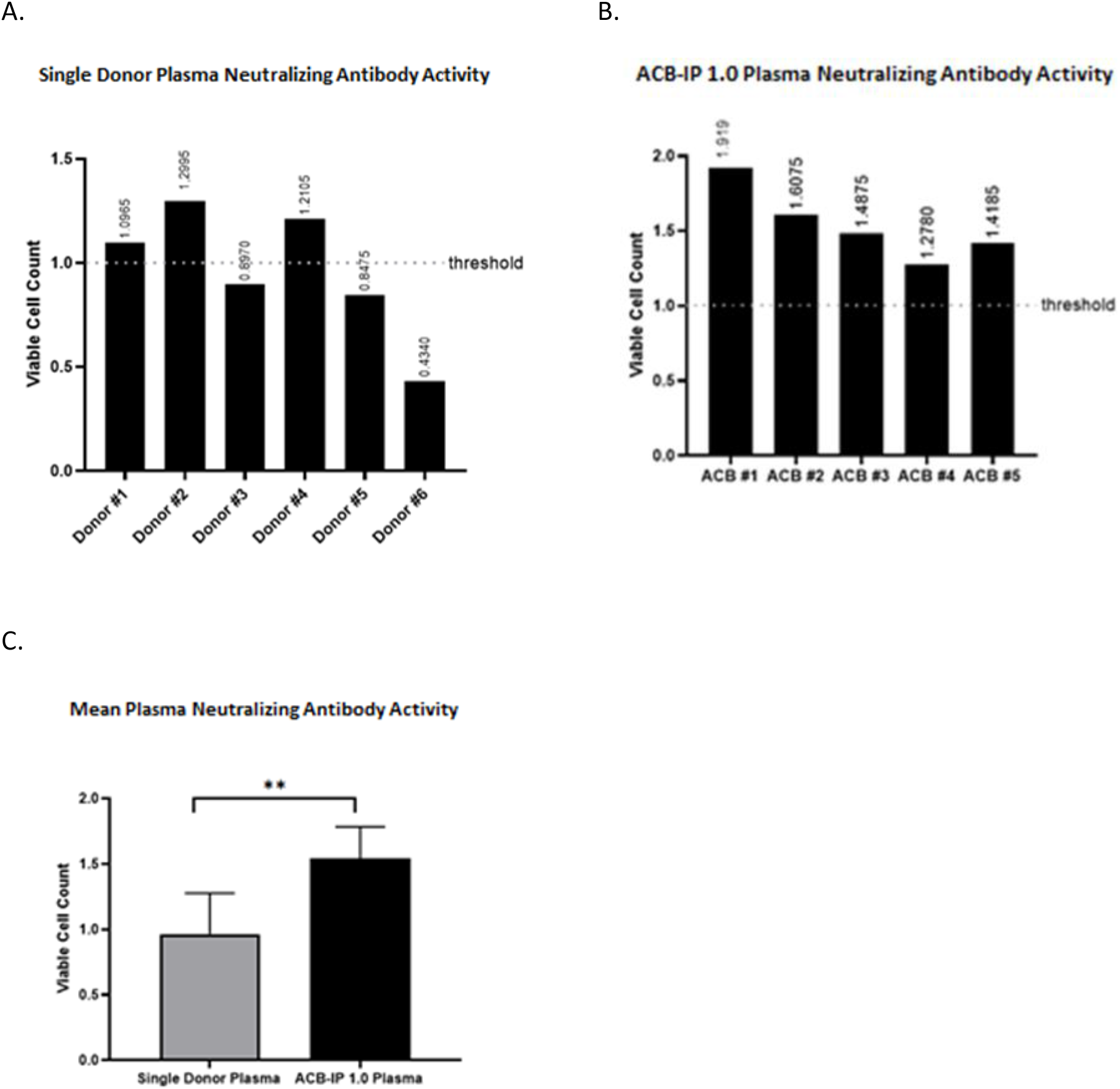
SARS-CoV2 Total Neutralizing Antibody Activity of Single Donor Plasma and ACB-IP 1.0 Plasma (in 1:256 dilution). A. Single donor plasma neutralizing antibody activity of each donors B. ACB-IP 1.0 plasma neutralizing antibody activity of each pooled product C. Comparison of mean neutralizing antibody activity **: ρ=0.0087

Total plasma neutralizing antibody activity between single donor plasma and ACB-IP 1.0 plasma showed no statistical significance (ρ= 0.93) in 1:128 dilution, while mean total plasma neutralizing antibody activity of ACB-IP 1.0 plasma (1.5421) was found statistically higher than single donor plasma (0.9642) in 1:256 dilution (ρ=0.0087) (Figure 2).

No correlation was found regarding SARS-CoV2 antibody titers and neutralizing antibody activity between both groups (single donor plasma antibody and neutralizing antibody activity correlation coefficient: 0.54, regression coefficient (R^2^): 0.44. ACB-IP 1.0 plasma antibody and neutralizing antibody activity correlation coefficient: −0.36, regression coefficient (R^2^): 0.79).

### Clinical Results

Clinical Characteristics of Single Donor plasma patients were presented in Table-1. Six out of nine single donor convalescent plasma patients were males. The median age of the patients was 65 years and none of them had a history of smoking. Two male patients had no coexisting diseases.

Clinical Characteristics of SARS-CoV-2-Infected Patients Who Received Single Donor Convalescent Plasma presented in Table-1.

| Patient | 1 | 2 | 3 | 4 | 5 | 6 | 7 | 8 | 9 |
| --- | --- | --- | --- | --- | --- | --- | --- | --- | --- |
| Sex | F | M | M | M | M | F | M | F | M |
| Age | 70s | 50s | 40s | 50s | 60s | 60s | 70s | 70s | 70s |
| Fever | No | No | Yes | Yes | Yes | No | Yes | Yes | Yes |
| Cough | No | Yes | Yes | Yes | Yes | No | Yes | No | Yes |
| Fatigue | Yes | Yes | Yes | Yes | Yes | No | Yes | Yes | Yes |
| Dyspnea | Yes | Yes | Yes | Yes | Yes | Yes | Yes | Yes | No |
| Diarrhea | No | Yes | No | No | No | No | No | No | No |
| Oxygen Supplement Requirement | Yes | Yes | Yes | Yes | No | Yes | Yes | Yes | No |
| Radiological Presentation | Multiple GGOs with consolidation | Multiple GGOs with consolidation | Multiple GGOs with consolidation | Multiple GGOs with consolidation, Bronchopneumonia | Multiple GGOs with consolidation, Partial Air Bronchogram | Multiple GGOs with consolidation, Atelectasis (Remains stable mostly - Decreasing) | Multiple GGOs with consolidation, Focal patched frosted dants | Density increase in heterogeneous structure compatible with consolidation | Multiple GGOs with consolidation, Interstitial septal thickening, Air cyst, Band atelectasis |
| Length of Stay In Hospital | 24 | 11 | 31 | 101 | 23 | 20 | 98 | 23 | 38 |
| Length Of Stay In ICU | 24 | 7 | 21 | 90 | 21 | 20 | 73 | 23 | 35 |
| Convalescent Plasma Application Date (Month) | May | April | April | April | April | April | April | May | April |
| Interval Between Disease On Set and Plasma Transfusion | 11 | 6 | 15 | 26 | 18 | 16 | 35 | 5 | 28 |
| Cycle of Convalescent Plasma | 1 | 1 | 1 | 2 | 1 | 1 | 1 | 1 | 1 |
| Convalescent Plasma Volume (ml) | 200 ml | 200 ml | 200 ml | 400 ml | 200 ml | 200 ml | 200 ml | 200 ml | 200 ml |
| Complications Prior Plasma Transfusion | No | No | No | No | No | No | No | No | No |
| Symptom Improvement | No | Yes | Yes | Yes | No | No | Yes | No | No |
| Radiological Improvement | Not be evaluated | Yes | No | Yes-Sequel Changes | No | Yes-Very little improvement | No | No | Not Evaluated |
| Current Status | Exitus | Discharged home | Discharged home | Discharged home | Exitus | Exitus | Discharged home | Exitus | Exitus |

Treatments of all patients were performed according to the Ministry of Health of Turkey Covid-19 treatment algorithms (Supplementary-2). Five patients received dornase-alpha and three patients received IL-6 blocker as SARS-CoV2 treatment.

One of the patients had two cycles of single donor convalescent plasma total of 400 ml volume. Volume loading due to transfusion was detected in one patient. No other reaction was observed.

The mean duration of the hospitalization stay was 41 days (11-101 days) and the mean duration of ICU stay was 34,9 days (7-90 days). After the treatments and convalescent plasma administration, four out of nine patients had clinical improvement and three patients had radiological improvement, one of the patients was not evaluated. In total, five patients were deceased.

Clinical characteristics of SARS-CoV2 infected patients who received pathogen-free, concentrated, pooled, convalescent plasma presented in Table-2. All of the patients treated with pathogen-free, concentrated, pooled convalescent plasma were males. The median age of the patients was 51 years and none of them had a history of smoking. Two of the patients had no coexisting diseases.

Characteristics of SARS-CoV-2-Infected Patients Who Received Pathogen-Free, Concentrated, Pooled, Convalescent Plasma presented in Table-2.

| Patient | 1 | 2 | 3 | 4 | 5 | 6 | 7 |
| --- | --- | --- | --- | --- | --- | --- | --- |
| Sex | Male | Male | Male | Male | Male | Male | Male |
| Age | 40s | 70s | 50s | 50s | 40s | 50s | 50s |
| Fever | Yes | No | No | Yes | Yes | Yes | No |
| Cough | Yes | Yes | No | Yes | Yes | No | Yes |
| Fatigue | No | Yes | No | Yes | Yes | Yes | Yes |
| Dyspnea | No | Yes | Yes | Yes | Yes | No | Yes |
| Diarrhea | No | Yes | No | No | No | Yes | No |
| Oxygen Supplement Requirement | No | Yes | Yes | Yes | No | No | Yes |
| Radiological Presentation | Multiple GGOs with consolidation | Multiple GGOs with consolidation | Multiple GGOs with consolidation and fibrosis streak - Patchy areas of GGOs | Multiple GGOs with consolidation | Multiple GGOs with consolidation and fibrosis streak | Multiple GGOs with consolidation | Multiple GGOs with consolidation and fibrosis streak |
| Length of Stay In Hospital | 12 | 172 | 13 | 19 | 24 | 5 | 61 |
| Length Of Stay In ICU | No | 139 | No | 10 | 24 | No | 37 |
| Convalescent Plasma Application Date (Month) | April | April | April | April | May | December | July |
| Cycle of Convalescent Plasma | 1 | 1 | 1 | 1 | 2 | 1 | 1 |
| Interval Between Disease On Set and Plasma Transfusion | 8 | 24 | 14 | 8 | 6 | 3 | 12 |
| Convalescent Plasma Volume (ml) | 144 | 156 | 170 | 136 | 142 & 144 | 200 | 176 |
| Complications Prior Plasma Transfusion | No | No | No | No | No | No | No |
| Symptom Improvement | Yes | Yes | Yes | Yes | Yes | Yes | Yes |
| Radiological Improvement | Yes | Yes | No | No | Yes | Yes | Yes |
| Current Status | Discharged home | Discharged home | Discharged home | Discharged home | Discharged home | Discharged home | Discharged home |

Treatments of all patients were performed according to the Ministry of Health of Turkey Covid-19 treatment algorithms. One patient received IL-6 blocker as SARS-CoV2 treatment. One of the patients had dornase-alpha as an add on treatment.

One of the patients had two cycles of pooled convalescent plasma total of 400 ml volume. No transfusion-related reactions had been observed. The mean duration of the hospitalization stay was 43,7 days (12-172 days) and the mean duration of ICU stay was 30 days (0-139 days).

After the treatments and convalescent plasma administration, four patients had clinical improvement and five patients had radiological improvement. A total of seven patients were discharged from the hospital.

There is no statistical difference between groups of patients that received single donor plasma and ACB-IP 1.0 plasma in concerning age (ρ=0.1604), radiological presentation (ρ= 0.999), co-existing disease (ρ= 0.999), oxygen supplement requirement (ρ= 0.5962).

In our study, we compare the two different patient groups regarding mortality rates and the length of hospitalization.

The mortality rate of the patients treated with ACB-IP 1.0 plasma showed statistically lower (p: 0,033) than the patients treated with single donor plasma. (Figure 3)

**Figure 3:**
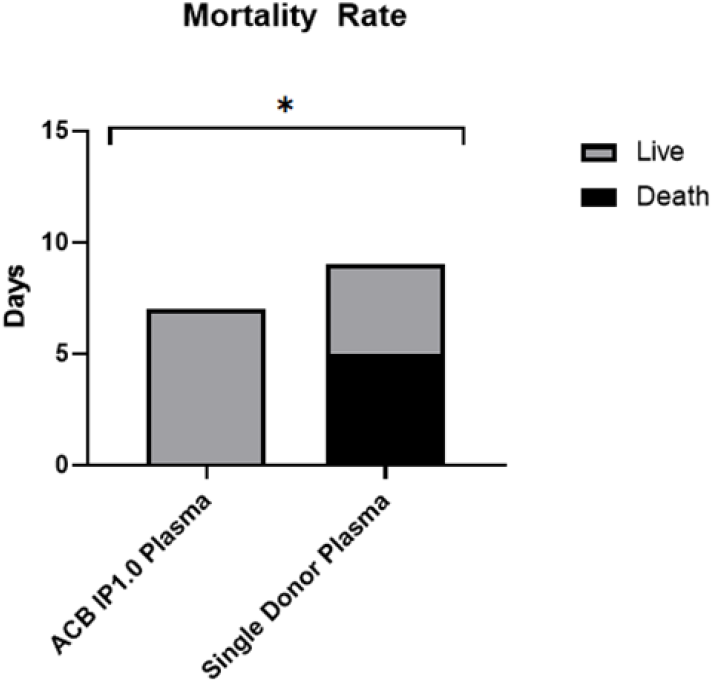
Comparison of the mortality rates between the two different plasma groups (*p:0,033)

The median length of hospital stay was 41 days for single donor plasma patients, and 43, 5 days for ACB-IP 1.0 plasma patients, showed no significant difference. The administration of either single donor plasma or ACB-IP 1.0 plasma to the patients within eight days significantly shortened the length of hospitalization compared to administration of either plasma to the patients later than eight days (ρ= 0,0021) (Figure 4).

**Figure 4:**
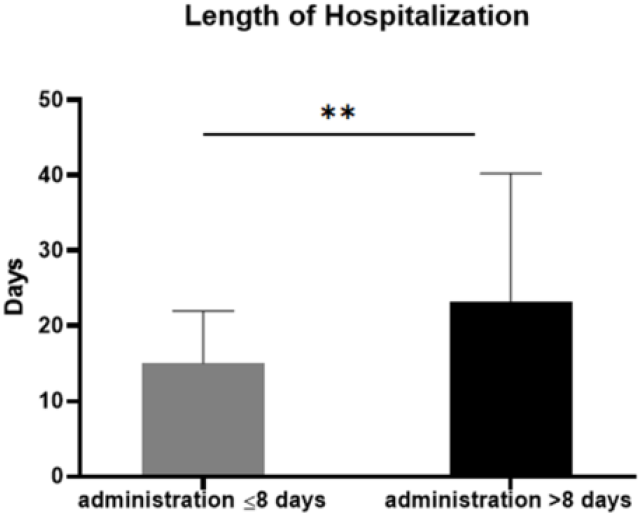
Effect of early plasma administration (8 days) on the length of hospitalization (*p:0,0021)

Although viral copy numbers could not be analyzed for all patients, only a single patient who received ACB-IP 1.0 had been analyzed. Three days after ACB-IP 1.0 was administered SARS-CoV2 antibody titer was above 25 log, and the viral copy number has decreased more than 20 log (Figure 5).

**Figure 5:**
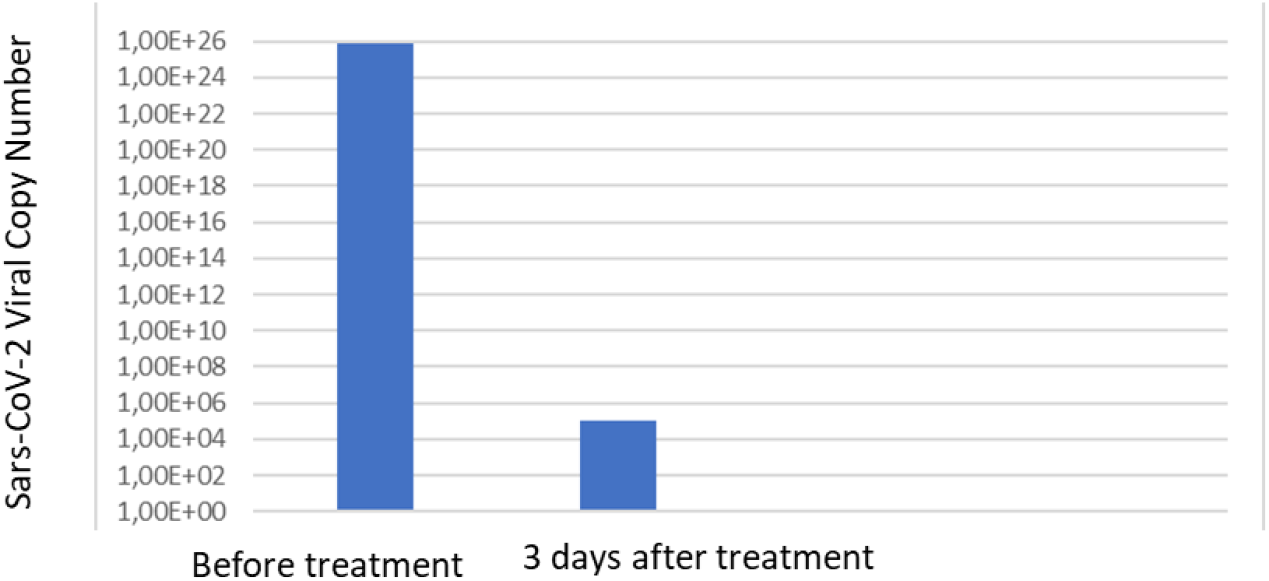
SARS-CoV2 Virus Copy Number of a Patient Who Received ACB-IP 1.0

## DISCUSSION

There are promising clinical trials regarding the beneficial effect of the convalescent plasma in SARS-CoV2 pneumonia. In the first published study involving five critically ill patients with COVID-19 and Acute Respiratory Distress Syndrome (ARDS), a single dose of convalescent plasma was administered, and clinical improvement had been achieved, despite an insufficient sample size.^18^ In another study, convalescent plasma of 200ml had been administered to ten patients with COVID-19 infection. It was shown that the patients were clinically improved and common laboratory parameters of infection such as decreased lymphocyte count and increased CRP tended to normalize.^13^ A recent prospective and propensity score-matched study, which compared the survival rates of convalescent plasma transfused 136 patients with non-transfused 251 patients, recommends administration of convalescent plasma within 72 hours of hospital admission due to observed significant reduction in mortality rates.^14^ In another study examined 20 severely and critically ill hospitalized COVID-19 patients and 20 matched controls.^16^ Although the study has some limitations such as short follow-up time and small sample size, concluded that convalescent plasma may improve survival if given early onset of the disease. In the study conducted by Liu et al. 39 hospitalized patients with severe to life-threatening COVID-19, received convalescent plasma compared to a non-transfused control group. According to their results patients receiving convalescent plasma therapy had improved survival and supplementary oxygen requirements at Day 14 post-transfusion compared to non-transfused controls. Moreover, the authors recommend the transfusion of the convalescent plasma immediately after the hospitalization which supports the current study results.^24^ In a large retrospective study it was revealed that convalescent plasma treatment is a safe treatment option in COVID-19.^25^ These findings are in line with our results that either plasma products are safe in clinical usage.

Conflicting with the above-mentioned studies, the beneficial effect of convalescent plasma could not be confirmed in SARS-CoV2 pneumonia patients in previous placebo-controlled trials.^17, 26^ The reason for the controversial result is explained with the lack of neutralizing antibody titer measurement of the donor plasma in the PLACID trial.^17^ Also, convalescent plasma was administered to the patients later than 3 days, contrary to what is recommended in the FDA report. FDA suggested in its report, that convalescent plasma transfusion can be effective within 3 days of COVID-19 diagnosis. In this report, FDA states that the signal to cut off ratio (s/co), which determines the amount of antibody in convalescent plasma, should be greater than 12 s/co, otherwise it should be healthcare providers may make an individualized assessment of benefit-risk to determine if the plasma unit is acceptable.^27^ The study conducted by V.A. Simonovich et al. it was shown that there was no relation between neither SARS-CoV2 antibody titers nor early plasma administration and clinical efficacy. However subsequent study of the same group, it had been demonstrated that early convalescent plasma administration of the older adults showed a significant reduction in the mortality rates.^28^

Since the total SARS-CoV2 antibody titers and neutralizing antibody titers of the single donor plasma showed inconsistency between donors, it was stated that this type of inconsistencies can affect clinical outcomes negatively.^17, 27^ In our study, SARS-CoV2 antibody titers and neutralizing antibody activities of the single donor plasma showed heterogeneity, s/c ratio demonstrated mostly below 12 s/co. However, ACB-IP 1.0 plasma which is a pooled concentrated plasma product showed higher SARS-CoV2 antibody titters and neutralizing antibody activities. As indicated there was no correlation between SARS-CoV2 antibody titers and neutralizing antibody activities in both single donor and ACB-IP 1.0 plasma. Donor #1 in the single donor plasma had 1,4 s/co as a total SARS-CoV2 antibody titer, but its neutralizing capacity showed sufficient result, similar to ACB-IP 1.0 pooled plasma product #1 had the lowest SARS-CoV2 antibody body titer (13.85 s/co) but showed the highest neutralizing capacity. Our study showed that ACB-IP 1.0 has more SARS-CoV2 antibody titer and neutralizing antibody activity which indicated mortality benefit compared to single donor plasma. Furthermore, the present study suggests that the early use of plasma showed a significant advantage in length of hospital stay; however statistical advantage in the early use of plasma on mortality rate was not shown, due to the small sample size.

Therefore, for a better understanding of the effectiveness of early convalescent plasma administration (within the first 72-hours of confirmed SARS-CoV2 infection) further studies should be performed among standardized plasma products.

However, prophylactic or early use may be dangerous in terms of transfusion-transmitted infections due to the individuals that do not meet the standard donor criteria. At this point, pathogen inactivation becomes a necessity. With the help of the pathogen inactivation process, the transfusion-transmitted infections are minimized and unsuitable individuals may be eligible. However, in our study, no data could be obtained to determine the advantages of pathogen inactivation due to its small scale.

This study has a potential limitation. It was not designed as a controlled-randomized study and the sample size is not enough to make a certain comment about the efficacy and safety of the ACB-IP 1.0 plasma product. Although the characteristics of patients were similar, the differences between co-therapies impair the comparison of plasma outcomes.

As a result, pooled pathogen inactive universal convalescent plasma may resolve the bias in SARS-CoV2 antibody titers and neutralizing antibody activities, without requiring blood group compatibility that allows patient accessibility in a shorter time and has safe plasma characteristic. This study indicates that ACB-IP 1.0 may be a superior product compared to standard single donor plasma.

## Data Availability

All data are available via Acibadem Hospital electronic medical records system.

## Acknowledgement/Disclaimers/Conflict of interest

Acıbadem Healthcare Group provides patients with gene therapy products and tissue engineering in the field of cellular therapy all of the 27 authors are affiliated with the funder.

## References

1. (WHO) WHO. Coronavirus Geneva WHO 2020 https://www.who.int/health-topics/coronavirus Access Date 21.1.2021. 2020.

2. Peiris JS, Yuen KY, Osterhaus AD, Stohr K. The severe acute respiratory syndrome. The New England journal of medicine. 2003; 349(25): 2431–41.

3. de Groot RJ, Baker SC, Baric RS, Brown CS, Drosten C, Enjuanes L, et al. Middle East respiratory syndrome coronavirus (MERS-CoV): Announcement of the Coronavirus Study Group. Journal of virology. 2013; 87(14): 7790–2.

4. Casadevall A, Pirofski LA. The convalescent sera option for containing COVID-19. The Journal of clinical investigation. 2020; 130(4): 1545–8.

5. Who Mers-Cov Research G. State of Knowledge and Data Gaps of Middle East Respiratory Syndrome Coronavirus (MERS-CoV) in Humans. PLoS currents. 2013; 5: ecurrents.outbreaks.0bf719e352e7478f8ad85fa30127ddb8.

6. Joyner MJ, Senefeld JW, Klassen SA, Mills JR, Johnson PW, Theel ES, et al. Effect of Convalescent Plasma on Mortality among Hospitalized Patients with COVID-19: Initial Three-Month Experience. medRxiv: the preprint server for health sciences. 2020: 2020.08.12.20169359.

7. Hung IFN, To KKW, Lee CK, Lee KL, Yan WW, Chan K, et al. Hyperimmune IV immunoglobulin treatment: a multicenter double-blind randomized controlled trial for patients with severe 2009 influenza A(H1N1) infection. Chest. 2013; 144(2): 464–73.

8. Stockman LJ, Bellamy R, Garner P. SARS: systematic review of treatment effects. PLoS medicine. 2006; 3(9): e343.

9. Hung IF, To KK, Lee CK, Lee KL, Chan K, Yan WW, et al. Convalescent plasma treatment reduced mortality in patients with severe pandemic influenza A (H1N1) 2009 virus infection. Clinical infectious diseases: an official publication of the Infectious Diseases Society of America. 2011; 52(4): 447–56.

10. Luke TC, Kilbane EM, Jackson JL, Hoffman SL. Meta-analysis: convalescent blood products for Spanish influenza pneumonia: a future H5N1 treatment? Annals of internal medicine. 2006; 145(8): 599–609.

11. Cheng Y, Wong R, Soo YO, Wong WS, Lee CK, Ng MH, et al. Use of convalescent plasma therapy in SARS patients in Hong Kong. European journal of clinical microbiology & infectious diseases: official publication of the European Society of Clinical Microbiology. 2005; 24(1): 44–6.

12. Ko JH, Seok H, Cho SY, Ha YE, Baek JY, Kim SH, et al. Challenges of convalescent plasma infusion therapy in Middle East respiratory coronavirus infection: a single centre experience. Antiviral therapy. 2018; 23(7): 617–22.

13. Duan K, Liu B, Li C, Zhang H, Yu T, Qu J, et al. Effectiveness of convalescent plasma therapy in severe COVID-19 patients. Proceedings of the National Academy of Sciences of the United States of America. 2020; 117(17): 9490–6.

14. Salazar E, Christensen PA, Graviss EA, Nguyen DT, Castillo B, Chen J, et al. Treatment of Coronavirus Disease 2019 Patients with Convalescent Plasma Reveals a Signal of Significantly Decreased Mortality. The American journal of pathology. 2020; 190(11): 2290–303.

15. Ahn JY, Sohn Y, Lee SH, Cho Y, Hyun JH, Baek YJ, et al. Use of Convalescent Plasma Therapy in Two COVID-19 Patients with Acute Respiratory Distress Syndrome in Korea: J Korean Med Sci. 2020 Apr 13;35(14):e149. doi: 10.3346/jkms.2020.35.e149.

16. Hegerova L, Gooley TA, Sweerus KA, Maree C, Bailey N, Bailey M, et al. Use of convalescent plasma in hospitalized patients with COVID-19: case series: Blood. 2020 Aug 6;136(6):759–762. doi: 10.1182/blood.2020006964.

17. Agarwal A, Mukherjee A, Kumar G, Chatterjee P, Bhatnagar T, Malhotra P. Convalescent plasma in the management of moderate covid-19 in adults in India: open label phase II multicentre randomised controlled trial (PLACID Trial). BMJ. 2020; 22(371).

18. Shen C, Wang Z, Zhao F, Yang Y, Li J, Yuan J, et al. Treatment of 5 Critically Ill Patients With COVID-19 With Convalescent Plasma. Jama. 2020; 323(16): 1582–9.

19. Hadjesfandiari N, Levin E, Serrano K, Yi Q-L, Devine DV. Risk analysis of transfusion of cryoprecipitate without consideration of ABO group. Transfusion. 2021; 61(1): 29–34.

20. Meliga SC, Farrugia W, Ramsland PA, Falconer RJ. Cold-induced precipitation of a monoclonal IgM: a negative activation enthalpy reaction. J Phys Chem B. 2013; 117(2): 490–4.

21. Middaugh CR, Gerber-Jenson B, Hurvitz A, Paluszek A, Scheffel C, Litman GW. Physicochemical characterization of six monoclonal cryoimmunoglobulins: possible basis for cold-dependent insolubility. Proceedings of the National Academy of Sciences of the United States of America. 1978; 75(7): 3440–4.

22. Josephson CD, Mullis NC, Van Demark C, Hillyer CD. Significant numbers of apheresis-derived group O platelet units have “high-titer” anti-A/A,B: implications for transfusion policy. Transfusion. 2004; 44(6): 805–8.

23. Nie J, Li Q, Wu J, Zhao C, Hao H, Liu H, et al. Quantification of SARS-CoV-2 neutralizing antibody by a pseudotyped virus-based assay. Nature protocols. 2020; 15(11): 3699–715.

24. Liu STH, Lin H-M, Baine I, Wajnberg A, Gumprecht JP, Rahman F, et al. Convalescent plasma treatment of severe COVID-19: a propensity score–matched control study. Nature medicine. 2020; 26(11): 1708–13.

25. Emery SL, Erdman DD, Bowen MD, Newton BR, Winchell JM, Meyer RF, et al. Real-time reverse transcription-polymerase chain reaction assay for SARS-associated coronavirus. Emerging infectious diseases. 2004; 10(2): 311–6.

26. Simonovich VA, Burgos Pratx LD, Scibona P, Beruto MV, Vallone MG, Vázquez C, et al. A Randomized Trial of Convalescent Plasma in Covid-19 Severe Pneumonia. New England Journal of Medicine. 2020.

27. DM. H. Emergency use authorization for COVID-19 convalescent plasma therapy. US Food and Drug Administration website. Published August 24, 2020. Accessed January 21, 2021. https://www.fda.gov/media/141477/download. 2020.

28. Joyner MJ, Wright RS, Fairweather D, Senefeld JW, Bruno KA, Klassen SA, et al. Early safety indicators of COVID-19 convalescent plasma in 5000 patients. The Journal of clinical investigation. 2020; 130(9): 4791–7.

